# Missense variants affecting the actin-binding domains of *PLS3* cause X-linked congenital diaphragmatic hernia and body wall defects

**DOI:** 10.1101/2021.07.07.21259278

**Authors:** Florence Petit, Mauro Longoni, Julie Wells, Richard Maser, Matthew J. Dysart, Hannah T.M. Contreras, Frederic Frénois, Eric Bogenschutz, Barbara R. Pober, Robin D. Clark, Philip F. Giampietro, Hilger H. Ropers, Hao Hu, Maria Loscertales, Xingbin Ai, Harrison Brand, Anne-Sophie Jourdain, Marie-Ange Delrue, Brigitte Gilbert-Dussardier, Louise Devisme, Boris Keren, David J. McCulley, Lu Qiao, Rebecca Hernan, Julia Wynn, Tiana M. Scott, Daniel G. Calame, Zeynep Coban-Akdemir, Patricia Hernandez, Andres Hernandez-Garcia, Hagith Yonath, James R. Lupski, Yufeng Shen, Wendy K. Chung, Daryl A. Scott, Carol J. Bult, Patricia K. Donahoe, Frances A. High

## Abstract

Congenital diaphragmatic hernia (CDH) is a relatively common and genetically heterogeneous structural birth defect associated with high mortality and morbidity. We describe eight unrelated families with a novel X-linked condition characterized by diaphragm defects, variable anterior body wall anomalies, and/or facial dysmorphism. Using linkage analysis and whole exome or whole genome sequencing, we identified novel missense variants in the actin binding domains of plastin 3 (*PLS3*), a gene encoding an actin bundling protein, that co-segregate with disease in all families. Loss-of-function variants in *PLS3* have been described previously in association with X-linked osteoporosis. To address these seemingly disparate clinical phenotypes, we performed *in silico* protein modeling and cellular overexpression experiments, which suggest that the affected residues in individuals with CDH are important for actin binding and result in disorganization of the actin cytoskeleton and a reduction in normal actin stress fiber formation. A mouse knock-in model of a variant identified in one of the families, p.W499C, shows partial perinatal lethality and recapitulates the key findings of the human phenotype, including diaphragm and abdominal wall defects. Both the mouse model and one surviving adult patient with a *PLS3* variant were observed to have increased, rather than decreased, bone mineral density. Together, these clinical and functional data in human and mouse reveal that specific missense variants affecting the actin binding domains of *PLS3* may have a gain-of-function effect and cause a new Mendelian disorder.

## INTRODUCTION

Congenital diaphragmatic hernia (CDH) is a common structural birth defect, occurring in 1:3,000 live births in the United States. Anatomically, CDH is characterized by incomplete formation or muscularization of the developing diaphragm, the most important respiratory muscle, together with lung hypoplasia. It is a severe birth defect, with estimated survival rates of 50-80%, despite advanced medical and surgical care. The most frequent and severe clinical complication is neonatal respiratory distress and hypertension of the pulmonary circulation, both of which can be refractory to standard treatments. Symptomatic infants with CDH require surgical intervention and often require extended stays in neonatal intensive care units and multiple invasive procedures including the use of extracorporeal membrane oxygenation. Among survivors, respiratory complications, neurodevelopmental deficits, and feeding difficulties requiring life-long medical attention are common ^1^.

The genetic causes of CDH, like other structural birth defects, are highly heterogeneous ^1 2^. The most frequently identified genetic causes of sporadic CDH are recurrent chromosome abnormalities ^1 2,3 4^ and deleterious *de novo* single gene variants ^5,6 7^ Familial CDH is thought to be rare because of the historically high morbidity and mortality in affected individuals, especially in syndromic forms, that impact reproductive fitness. However, studies of rare familial clusters have revealed important human CDH genes ^8,9 10^. Several lines of evidence suggest X-linked loci for CDH: kindreds showing a pattern consistent with X-linked inheritance, a slightly increased M:F ratio among affected patients^11,12^, and the occurrence of diaphragmatic defects in several X-linked syndromic conditions ^1 2^.

In this report, we describe eight unrelated families with novel variants in the X-linked gene *PLS3*. The families, including five families with multiple affected individuals, show transmission of congenital diaphragm defects compatible with X-linked inheritance. Sequencing studies identified inherited or *de novo* missense variants in *PLS3* that segregate with affected status in all eight families. *PLS3* maps to Xq23 and encodes plastin 3 (also known as T-plastin), an actin-bundling protein. Loss of function (LOF) alleles and deletions in *PLS3* have been reported previously in families with X-linked osteoporosis and osteoporotic fractures (MIM# 300910). Affected males present with childhood osteoporosis and fractures of the axial and appendicular skeleton, while the bone phenotype in affected females ranges from normal bone density to early-onset osteoporosis ^13^.

The actin cytoskeleton is a dynamic network that is important for multiple cellular processes in eukaryotes. Actin structures have distinct intracellular localization and are bound to different actin cross-linking proteins, among which are actin-bundling proteins of the plastin family^14,15^. *PLS3* encodes one of the three vertebrate Plastin proteins, which bundle together adjacent actin filaments in a calcium-responsive manner. Plastin proteins contain an N-terminal calcium-binding region and two C-terminal actin binding domains that are each comprised of two calponin homology domains. Each actin binding domain binds to adjacent actin filaments, thereby linking them into bundles^16^. These tight actin bundles are critical for formation of specialized actin structures such as stress fibers, filopodia, lamellipodia, and microvilli ^15^.

The CDH-associated *PLS3* variants all affect residues in the actin binding domains of the protein. These variants are predicted to alter actin binding without causing major conformational changes in the protein in silico, and result in abnormal actin structures in a cell culture model. We also generated a mouse model carrying a humanized sequence for one of the variants observed in patients in our study (p.W499C) that showed variable perinatal lethality, diaphragm and abdominal wall defects, and increased bone mineral density. These data support a novel pathophysiologic mechanism leading to CDH and/or anterior body wall anomalies with nearly complete penetrance in male probands and variable expressivity in female carriers.

## MATERIAL AND METHODS

### Patient recruitment

The study participants were recruited and provided written informed consent according to clinical investigation standards of the Partners/Mass General Brigham Human Research Committee, Boston Children’s Hospital Institutional Review Board, Columbia University Irving Medical Center (CUIMC) Institutional Review Board (IRB), and the Baylor College of Medicine Institutional Review Board. All IRB protocols were approved. Written informed consent was obtained from all study participants in accordance with the local IRB protocol and specific consent for photography was obtained where applicable. Collaboration between investigators was aided by the GeneMatcher community^17^. Clinical and research microarrays were performed on probands and excluded causative copy number variants in affected individuals (data not shown). Detailed clinical data collection was performed by chart review and, where possible, physical examination by members of the study staff.

### DNA extraction

DNA was extracted from peripheral blood samples in EDTA using the DNeasy Blood & Tissue Kit (Qiagen) according to the accompanying handbook. Formalin-fixed and paraffin-embedded (FFPE) tissue was processed with QIAamp DNA FFPE Tissue Kit (Qiagen). Extracted DNA was quantitated with the Quant-iT PicoGreen dsDNA Assay Kit (Thermo Fisher Scientific).

### Linkage analysis

Single nucleotide polymorphism (SNP)-data were collected on members of family 1 using the Infinium HumanOmniExpress-24 v1.0 (Illumina) platform. Parametric linkage analysis for X-linked traits was carried out using MINX in the Merlin software package^18 18^, assuming a disease allele with frequency 0.0001 in our model.

### Next-generation sequencing

Whole exome sequencing (WES) for families 2-4 was performed at the University of Washington Department of Genome Sciences as described previously^6^. WES for families 7 and 8 were performed at Baylor College of Medicine Human Genome Sequencing Center using an Illumina dual indexed, paired-end pre-capture library per manufacturer protocol with modifications (https://www.hgsc.bcm.edu/content/protocols-sequencing-library-construction). Libraries were pooled and hybridized to the HGSC VCRome 2.1 plus custom Spike-In design according to the manufacturer’s protocol (NimbleGen) with minor revisions^19^. Paired-end sequencing was performed with the Illumina NovaSeq6000 platform. The sample achieved 98% of the targeted exome bases covered to a depth of 20x or greater and had a sequencing yield of 13.2 Gb. Illumina sequence analysis was performed using the HGSC HgV analysis pipeline which moves data through various analysis tools from the initial sequence generation on the instrument to annotated variant calls (SNPs and intra-read in/dels) ^20,21^. In parallel to the exome workflow a SNP Trace panel was generated for a final quality assessment. This included orthogonal confirmation of sample identity and purity using the Error Rate In Sequencing (ERIS) pipeline developed at the BCM-HGSC. Using an “e-GenoTyping” approach, ERIS screens all sequence reads for exact matches to probe sequences defined by the variant and position of interest. A successfully sequenced sample must meet quality control metrics of ERIS SNP array concordance (>90%) and ERIS average contamination rate (<5%). WGS for families 5 and 6 was processed at the Broad Institute Genomic Services as described previously^22^.

WES and WGS data were processed using GATK Best Practice v4.0. using pipelines described previously ^6,7,22^. WES interpretation was performed with Seqr (seqr.broadinstitute.org) and/or GEMINI: Integrative Exploration of Genetic Variation and Genome Annotations ^23^. The following filters were used for analysis of both WES and WGS: quality filters GQ>20, AB>25, coding variants with predicted moderate to high impact including loss-of-function variants (frameshift, nonsense, essential splice site, in-frame indel) and missense variants, allele frequency below 0.01% in the control population database gnomAD, including any of their subpopulations^24^. All rare missense variants were reviewed individually regardless of algorithm predictions of deleterious effect. All reported variants were visualized manually using Integrative Genome Viewer (IGV; http://software.broadinstitute.org/software/igv) and validated by Sanger sequencing.

#### Data sharing

WES data from families 2-4 has been deposited into the NIH National Center for Biotechnology Information (NCBI) Database of Genome & Phenotypes (dbGAP). WES data from families 7 and 8 will be deposited into NHGRI Genomic Data Science Analysis, Visualization, and Informatics Lab-space (AnVIL). WGS data for families 5 and 6 will be deposited into the data repository of the NICHD Gabriella Miller Kid’s First (GMKF) Program.

### Protein modeling

Structural modeling is based on the N-terminal actin-crosslinking domain structure from human plastin 3 obtained by X-ray diffraction(1AOA) ^25^, on the actin-crosslinking core of Arabidopsis fimbrin (1PXY) by X-ray diffraction^26^, and on the fourth CH domain from human plastin 3 T-isoform (IWJC) by MNR (PDB ID: 1WJO) [unpublished data by deposition authors: Tomizawa, T., Kigawa, T., Koshiba, S., Inoue, M., Yokoyama, S., RIKEN Structural Genomics/Proteomics Initiative (RSGI)]. Data files can be downloaded from the Protein Data Bank (PDB, www.rcsb.org).

### Overexpression of *PLS3* variants

Recombinant human *PLS3* constructs were generated using the high copy number pRP[Exp]-CMV>HA/hPLS3[NM_005032.6] expression vector under the control of the constitutively active CMV promoter (VectorBuilder Inc (Santa Clara, CA)). A hemoagglutinin (HA) tag was fused to the N-terminus of the PLS3 protein for the purpose of immunofluorescence. In situ mutagenesis was performed with the Q5® Site-Directed Mutagenesis Kit (New England Biolabs, Ipswich, MA) to introduce patient-specific variants, and the resulting plasmids were transformed into One Shot TOP10 chemically competent E. coli cell (Invitrogen). Mutagenesis primers were designed using the NEBaseChanger v.1.2.8 (nebasechanger.neb.com) (vector map, primer sequences, and amplification conditions are provided as Supplemental Materials). Successful mutagenesis was confirmed by Sanger sequencing. Plasmid purification was performed with the QIAGEN Plasmid Mini Kit for colony screening or with the QIAGEN Plasmid Maxi Plus Kit (QIAGEN Inc., Germantown, MD) for transient transfection experiments. RFL-6 cells (ATCC® CCL-192^™^) were cultured in modified Ham’s F-12 Medium with 1% L-glutamine (Corning Inc., Corning, NY), supplemented with 10% fetal bovine serum and 1% Penicillin-Streptomycin. Plasmid DNA (500 ng) was electroporated into 100,000 cells using the Neon® Transfection System (Invitrogen^™^, Carlsbad, CA) with a single 1650 V and 20 ms pulse. Electroporated cells were plated and maintained in Opti-MEM^™^ Reduced Serum Medium for 24 hours on coverslips in a 24-well plate until fixation with 4% paraformaldehyde and staining. The complete list of antibodies and dyes is provided as Supplemental Materials. Electroporation experiments were conducted at least in triplicate and efficiency monitored via parallel transfection of a green fluorescent protein (GFP) expressing plasmid. Images were collected with a Nikon Eclipse 80i equipped with a Photometrics CoolSNAP DYNO Scientific CCD Camera and the NIS-Elements BR 5.02.01 64-bit softwareri. Quantification of actin stress fibers was performed in quadruplicate for each condition, and for each replicate 61-145 images of randomly selected transfected cells were analyzed. A subjective assessment of normal vs. abnormal (reduced) actin stress fibers in each cell was performed by two independent observers who were blinded to the experimental condition. Data are presented per condition as an average of the percentage of cells across replicates displaying normal actin stress fibers, and statistical significances between conditions were calculated using a two-tailed t test.

### Whole mount In situ hybridization

The mouse *Pls3* probe was generated using a 734 segment of the mouse transcript that was PCR amplified (PCR Master Mix, Promega, Madison, WI) with one set of exon-exon boundary overlapping primers (forward: gagctagcagcgtaggtcg and reverse: cattttgcagagcacgatccc). The purified PCR fragment was cloned into the pCR^™^II-TOPO® TA vector (TOPO® TA Cloning® Kit, Dual Promoter) (ThermoFisher, Life Technologies Corporation), and transformed into One Shot® TOP10 Chemically Competent Cells (ThermoFisher, Life Technologies Corporation). Sense and anti-sense Digoxigenin-11-UTP labeled probes (DIG RNA Labeling Mix, Sigma-Aldrich) were synthesized with SP6 and T7 RNA polymerases, respectively. E12.5 mouse embryos were collected in cold phosphate-buffered saline (PBS), fixed over night at 4°C with 4 % paraformalehyde in PBS, and washed with PBT (PBS with 0.1% Tween 20) at 4°C. Whole-mount in situ hybridization was performed as described^27^. Embryos were bleached with 6% hydrogen peroxide in PBT for 1 hour, treated with 10ug/mL Proteinase K (Sigma-Aldrich) in PBT for 10 minutes, then incubated in pre-warmed hybridization buffer at 70°C for 1 hour. DIG-labelled probe was added to fresh hybridization buffer at a concentration of 1ug/mL, and dissected embryos were incubated in this buffer at 70°C overnight. Embryos were then washed stringently and incubated overnight at 4°C in preabsorbed 1:2000 anti-DIG antibody (Roche) with 0.1% goat serum in TBST. Probe was visualized by incubating dissected embryos for 24-48 hours in BM-Purple (Roche).

Embryos were then further dissected to isolate the lung for imaging using a Nikon AZ100 microscope.

### *Pls3* mouse model generation

All research involving animals complied with protocols approved by the Institutional Animal Care and Use Committees (IACUC) from The Jackson Laboratory and Massachusetts General Hospital. *Pls3* mice, containing either the W499C substitution in (*Pls3*^*em1Bult*^, referred to as *Pls3*^*W499C*^) or a 14 bp deletion (*Pls3*^*em2Bult*^, referred to as *Pls3*^*14bpdel*^) were created at The Jackson Laboratory. To introduce the *Pls3* W499C variant into the mouse genome, *S. pyogenes* Cas9 gene editing reagents were designed using the Benchling online software tool (benchling.com) targeting *Pls3* tryptophan codon 499 (transcript ID *Pls3-*201, ENMUST00000033547). Introduction of a C->G variant at mouse chrX:75793603 (positive strand, GRCm38/mm10) results in a change in codon from tryptophan (TGG) to cysteine (TGC) on the negative strand. A single guide RNA (gRNA) sequence (CCTTGACCTTGGCTGTAGTC) was ordered as an ALT-R crRNA and hybridized with ALT-R tracRNA (IDT, Research Triangle Park, NC). A single-stranded oligonucleotide donor to introduce the W499C variant (variant underlined, 5’ AATATACTAAGGGTGAATTCCTATATGCATATTACTCTGACGCTTCCATAGCCAATACTGTGCACGAT ACCTTCTCATCAGCTG<u>G</u>CAGACTACAGCCAAGGTCAAGGTAGGGTTGCCATCGTTCAGGT-3’) was synthesized as an Extremer by Eurofins (USA). Fertilized mouse embryos were generated by natural mating following superovulation of C57BL/6J females and cultured as described previously^28^. Guide RNA, Cas9 protein (IDT), and donor oligos were introduced into C57BL/6J single cell zygotes by electroporation as described^28^. Manipulated embryos were immediately transferred into pseudopregnant female mice in order to obtain liveborn pups. *Pls3*^*W499C*^ and *Pls3*^*14bpdel*^ mice were obtained from the same electroporation experiment as a consequence of different zygotes utilizing different mechanisms of DNA repair.

Pls3 mice were genotyped by PCR using the primers listed below. DNA was isolated from tissues obtained by either ear notching or tail tipping using the Hot Shot method (Truett GE et al. Biotechniques 2000 Jul:29(1):52-54). Each 20 ml PCR reaction consisted of 1 ml of DNA, 1 ml of each genotyping primer, 4 ml of 5M betaine (Sigma-Aldrich, St. Louis, MO), 4 ml of 5X Phusion buffer (New England Biolabs, Beverly, MA), 2 ml of dNTP mix (10 mM each dNTP; Promega, Madison, WI) and 0.2 ml of Phusion DNA polymerase (New England Biolabs, Beverly, MA). Reactions were amplified by 28 cycles of PCR with an annealing temperature of 61°C and visualized by electrophoresis on a 1.8% agarose gel. The wild type and KI PCR products are 608 base pairs (bp) in length while the 14 bp del PCR product is 594 bp. The following primers were used: Pls3 9064 5’-AGGGAACTCCATGAGAACATCTG-3’and Pls3 9062 5’-GCTTCTGGAGGAAAGAACTAGATC-3’. For animals from the Pls3 colonies, PCR reactions were purified using MagBio High Prep PCR magnetic beads (Gaithersburg, MD) following manufacturer’s protocol and subsequently analyzed by Sanger sequencing using primer 9064.

### Mouse diaphragm dissections

Mice were collected at E18.5 (embryos) or P0 (neonates), fixed overnight in 4% PFA at 4°C, and dissected for gross visualization of the body wall, diaphragm, and lungs. Photographs were taken using a Nikon AZ100 microscope. Diaphragm phenotypes were scored from photographs of E18.5 or P0 diaphragms and the investigator was blinded to the genotypes of the animals during the data collection.

### Mouse whole mount staining of the body wall

Postnatal day 0 pups were euthanized through decapitation, the skin along the spine was cut open, and bodies were fixed overnight in 4% paraformaldehyde at 4°C. The fixed bodies were then skinned under a dissection microscope in 1x PBS. Samples were bleached for 2 hours at room temperature in Dent’s Bleach (1:2 30% H_2_O_2_:Dent’s fix), washed three times in Methanol then stored for 2 weeks in Dent’s fix (1:4 DMSO:methanol) at 4°C. Samples were then washed in PBS, incubated for 1 hour in PBS at 65°C, blocked for 1 hour in 5% serum and 20% DMSO, then incubated in alkaline phosphatase–conjugated mouse IgG1 antibody to My32 (Sigma #A4335) for 48 hours. Staining was then detected using a 1:1 NTMT:NBT/BCIP Substrate Solution (ThermoFisher #34042) for 40 minutes at room temperature. Images were taken on a Leica M125 stereo microscope. Distance between stained oblique muscles were measured using Adobe Photoshop. Mean distance between stained oblique muscles were compared using Welch’s t test.

### Mouse bone densitometry

Wild type C57BL/6J (The Jackson Laboratory, Bar Harbor, ME stock 000664), *Pls3*^*W499C*^ KI and *Pls3*^*14bpdel*^ mice were analyzed by dual energy x-ray absorptiometry (DEXA) at 3 months of age using an UltraFocus scanner (Faxitron, Tuscon, AZ). For this procedure, mice were weighed and anesthetized with continuous inhalation of isofluorane and four sets of images were acquired at low and high energy. Collected data was analyzed by VisionDXA software (using body minus head protocol) and reported as total body weight (g), total tissue mass (g), lean tissue mass (g), fat mass (g), percentage of body weight as fat, total body area (cm^2^), total bone area (cm^2^), bone mineral content (g) and bone mineral density (g/cm^2^). Eight wildtype females, 8 wildtype males, 7 *Pls3*^*14bpdel/14bpdel*^ homozygous females, 8 *Pls3*^*14bpdel*^ hemizygous males, 6 *Pls3*^*W99C/W499C*^ homozygous females, and 3 *Pls3*^*W499C*^ hemizygous males were studied.

## RESULTS

### Clinical Case Descriptions

The families were recruited individually into separate studies at the University of Lille, Boston Children’s Hospital and Massachusetts General Hospital CDH genetics study, Diaphragmatic Hernia Research & Exploration, Advancing Molecular Science (DHREAMS), and Baylor College of Medicine. Collaborations were facilitated through GeneMatcher^17^ and connections between researchers across CDH genetics study cohorts. Pedigrees are shown in Figure 1 and clinical details are provided in Table 1.

**FIGURE 1.** Pedigrees have been removed from the preprint version of the manuscript. Please contact the corresponding author for additional information. (Pedigrees removed from pre-print version of manuscript)

**TABLE 1:**
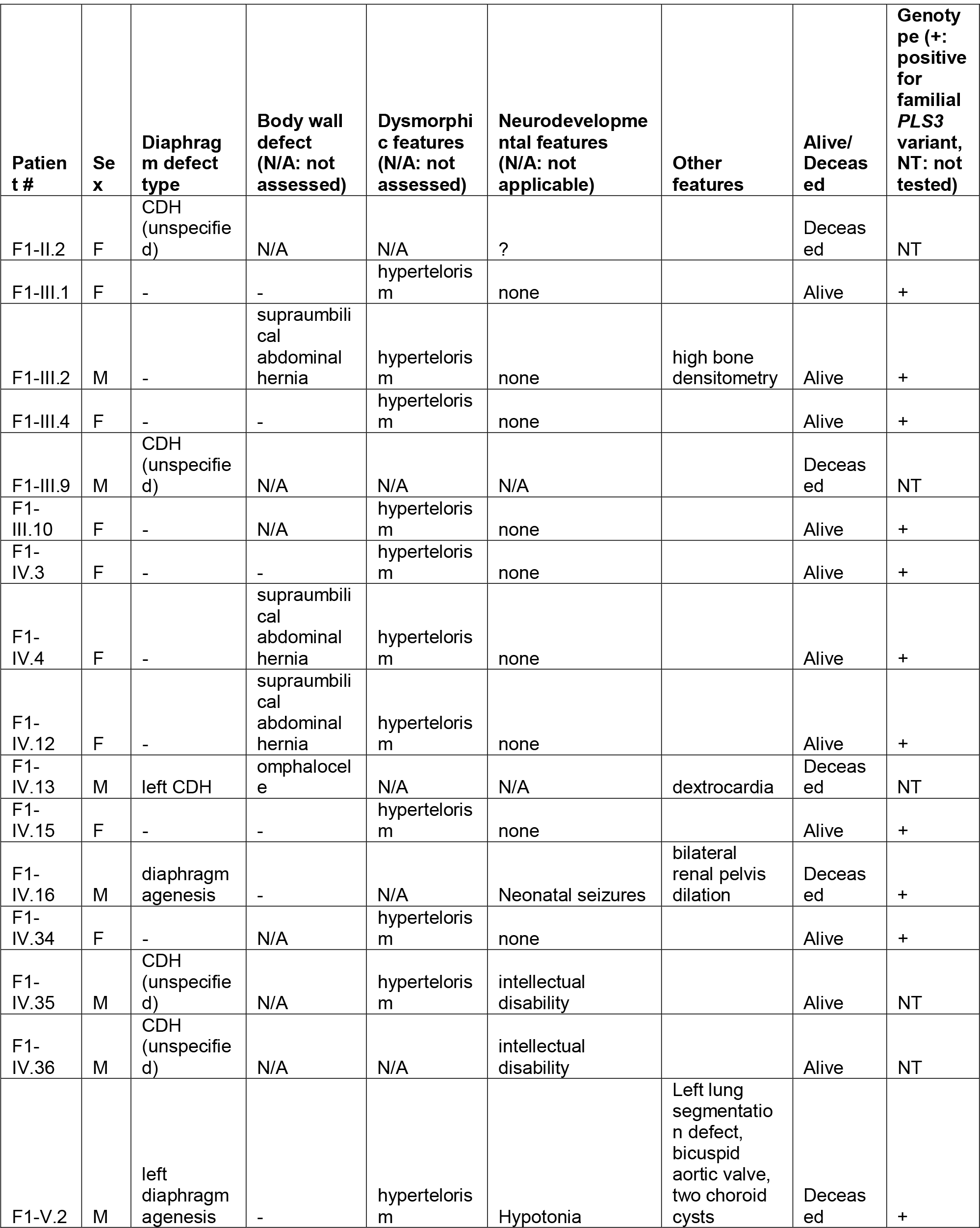
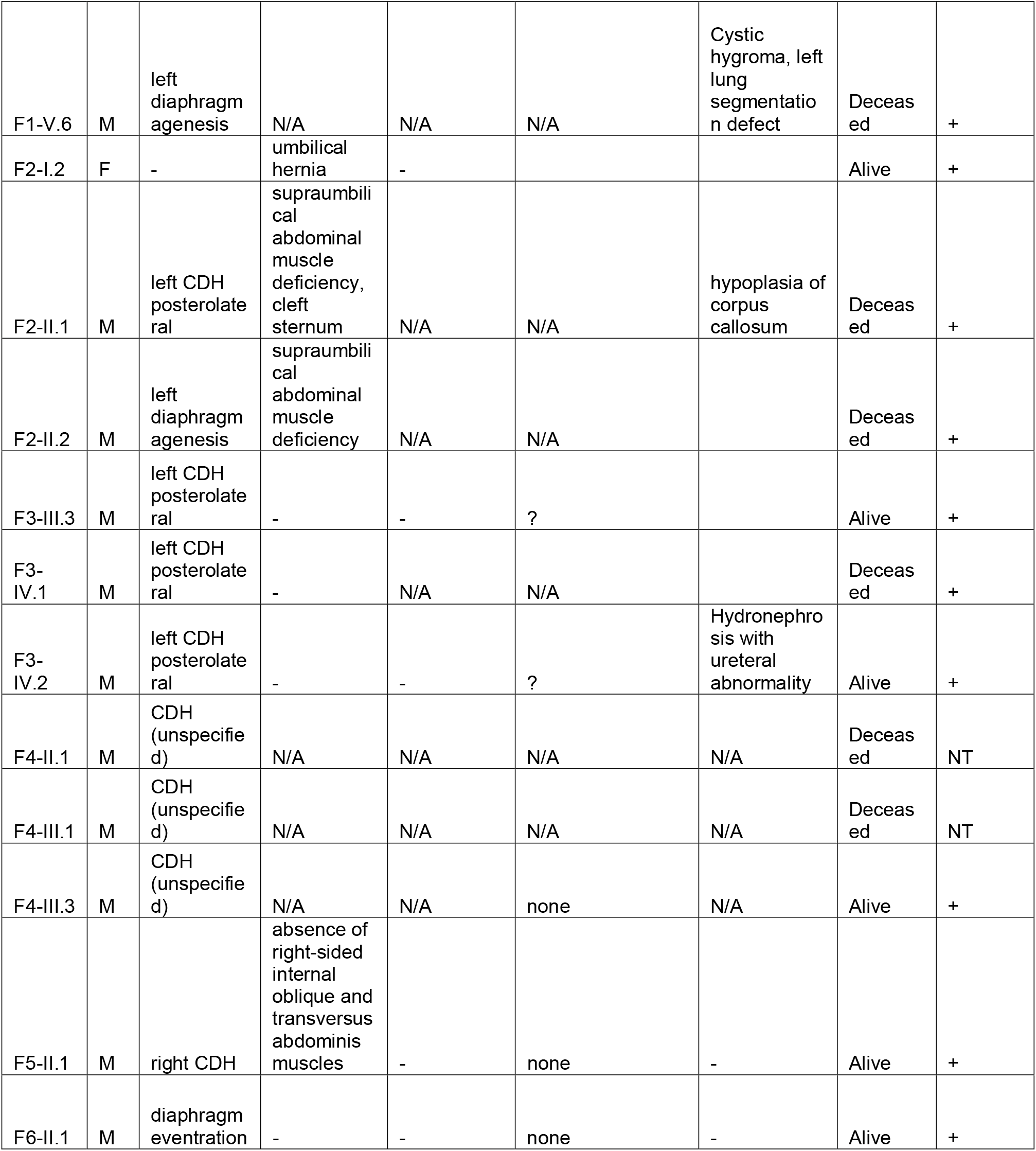
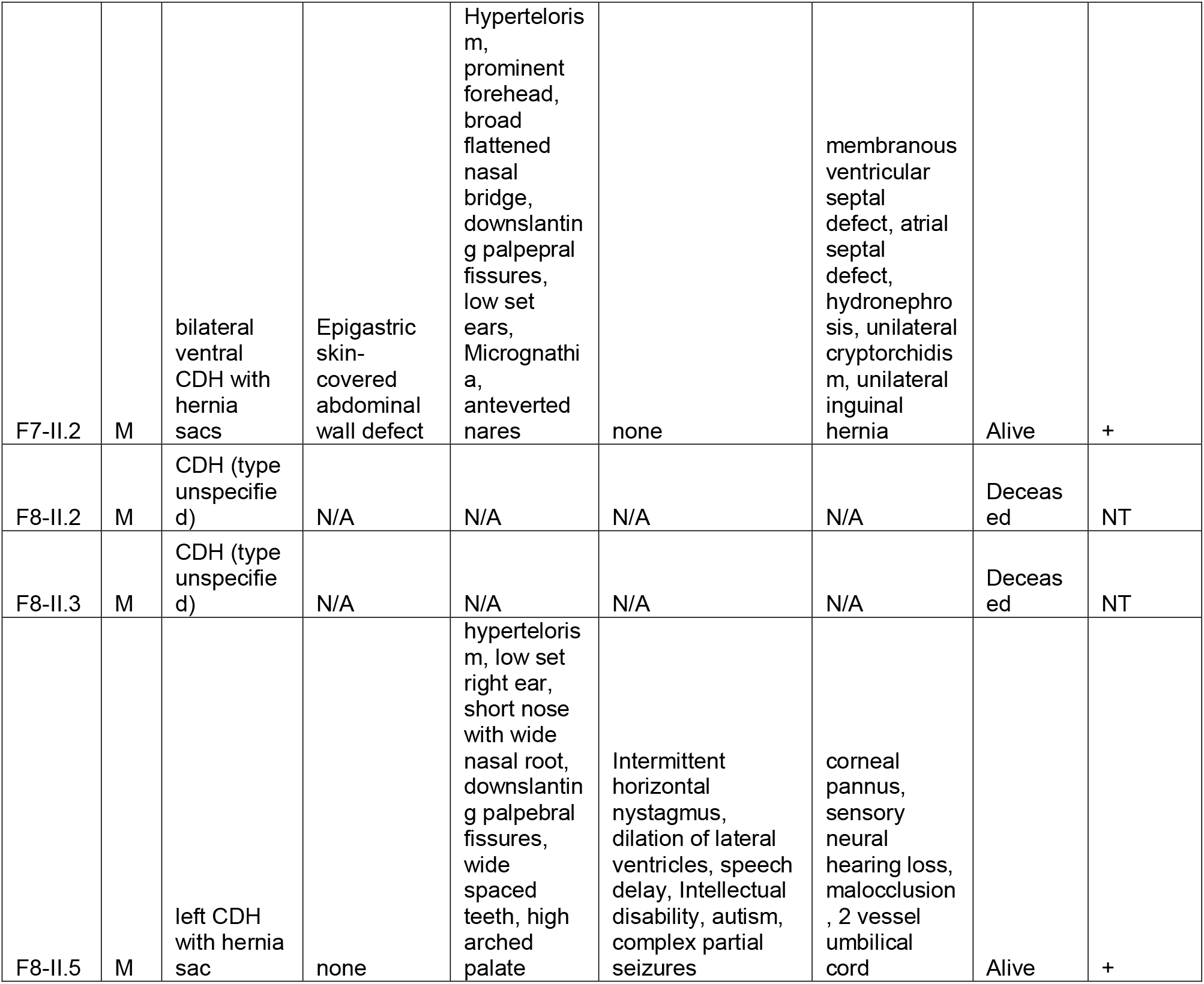
Clinical information for affected individuals.

| Patient # | Sex | Diaphragm defect type | Body wall defect (N/A: not assessed) | Dysmorphic features (N/A: not assessed) | Neurodevelopmental features (N/A: not applicable) | Other features | Alive/Deceased | Genotype (+: positive for familial PLS3 variant, NT: not tested) |
| --- | --- | --- | --- | --- | --- | --- | --- | --- |
| F1-II.2 | F | CDH (unspecified) | N/A | N/A | ? |  | Deceased | NT |
| F1-III.1 | F | - | - | hypertelorism | none |  | Alive | + |
| F1-III.2 | M | - | supraumbilical abdominal hernia | hypertelorism | none | high bone densitometry | Alive | + |
| F1-III.4 | F | - | - | hypertelorism | none |  | Alive | + |
| F1-III.9 | M | CDH (unspecified) | N/A | N/A | N/A |  | Deceased | NT |
| F1-III.10 | F | - | N/A | hypertelorism | none |  | Alive | + |
| F1-IV.3 | F | - | - | hypertelorism | none |  | Alive | + |
| F1-IV.4 | F | - | supraumbilical abdominal hernia | hypertelorism | none |  | Alive | + |
| F1-IV.12 | F | - | supraumbilical abdominal hernia | hypertelorism | none |  | Alive | + |
| F1-IV.13 | M | left CDH | omphalocele | N/A | N/A | dextrocardia | Deceased | NT |
| F1-IV.15 | F | - | - | hypertelorism | none |  | Alive | + |
| F1-IV.16 | M | diaphragm agenesis | - | N/A | Neonatal seizures | bilateral renal pelvis dilation | Deceased | + |
| F1-IV.34 | F | - | N/A | hypertelorism | none |  | Alive | + |
| F1-IV.35 | M | CDH (unspecified) | N/A | hypertelorism | intellectual disability |  | Alive | NT |
| F1-IV.36 | M | CDH (unspecified) | N/A | N/A | intellectual disability |  | Alive | NT |
| F1-V.2 | M | left diaphragm agenesis | - | hypertelorism | Hypotonia | Left lung segmentation defect, bicuspid aortic valve, two choroid cysts | Deceased | + |
| F1-V.6 | M | left diaphragm agenesis | N/A | N/A | N/A | Cystic hygroma, left lung segmentation defect | Deceased | + |
| F2-I.2 | F | - | umbilical hernia | - |  |  | Alive | + |
| F2-II.1 | M | left CDH posterolateral | supraumbilical abdominal muscle deficiency, cleft sternum | N/A | N/A | hypoplasia of corpus callosum | Deceased | + |
| F2-II.2 | M | left diaphragm agenesis | supraumbilical abdominal muscle deficiency | N/A | N/A |  | Deceased | + |
| F3-III.3 | M | left CDH posterolateral | - | - | ? |  | Alive | + |
| F3-IV.1 | M | left CDH posterolateral | - | N/A | N/A |  | Deceased | + |
| F3-IV.2 | M | left CDH posterolateral | - | - | ? | Hydronephrosis with ureteral abnormality | Alive | + |
| F4-II.1 | M | CDH (unspecified) | N/A | N/A | N/A | N/A | Deceased | NT |
| F4-III.1 | M | CDH (unspecified) | N/A | N/A | N/A | N/A | Deceased | NT |
| F4-III.3 | M | CDH (unspecified) | N/A | N/A | none | N/A | Alive | + |
| F5-II.1 | M | right CDH | absence of right-sided internal oblique and transversus abdominis muscles | - | none | - | Alive | + |
| F6-II.1 | M | diaphragm eventration | - | - | none | - | Alive | + |
| F7-II.2 | M | bilateral ventral CDH with hernia sacs | Epigastric skin-covered abdominal wall defect | Hypertelorism, prominent forehead, broad flattened nasal bridge, downslanting palpebral fissures, low set ears, Micrognathia, anteverted nares | none | membranous ventricular septal defect, atrial septal defect, hydronephrosis, unilateral cryptorchidism, unilateral inguinal hernia | Alive | + |
| F8-II.2 | M | CDH (type unspecified) | N/A | N/A | N/A | N/A | Deceased | NT |
| F8-II.3 | M | CDH (type unspecified) | N/A | N/A | N/A | N/A | Deceased | NT |
| F8-II.5 | M | left CDH with hernia sac | none | hypertelorism, low set right ear, short nose with wide nasal root, downslanting palpebral fissures, wide spaced teeth, high arched palate | Intermittent horizontal nystagmus, dilation of lateral ventricles, speech delay, Intellectual disability, autism, complex partial seizures | corneal pannus, sensory neural hearing loss, malocclusion, 2 vessel umbilical cord | Alive | + |

**Family 1** is a large family including 8 affected males and several more mildly affected females. Most affected males died because of diaphragm defects; however, three affected males survived to adulthood. Additionally, one female family member (F1-II.2) presented with a mild form of CDH that was surgically repaired. All surviving affected males and carrier females were noted to have distinct facial features, which include hypertelorism (Data not shown). Only one affected male from this family did not present with CDH (F1-III.2) but had an extensive supraumbilical abdominal hernia that was surgically repaired shortly after birth (Data not shown). Supraumbilical hernia was also a frequent finding in carrier females (Data not shown). Among the three males that survived to adulthood, one had no neurodevelopmental findings, though two of the affected males with CDH had intellectual disability (F1-IV.35 and F1-IV.36). In both cases the possibility that the intellectual disability was related to neonatal hypoxia-ischemia could not be ruled out.

As males with loss-of-function variants in *PLS3* have been described to have severe and early-onset osteoporosis^13^, we investigated bone mineralization in one proband of family 1 (F1-III.2) with dual-energy x-ray absorptiometry (DEXA). DEXA measurements on the left hip (Z score +2, T score +1.2) and lumbar vertebrae (Z score +5.8, T score +5.4) showed instead increased bone mineral density in this individual (Figure 2C).

**FIGURE 2.**
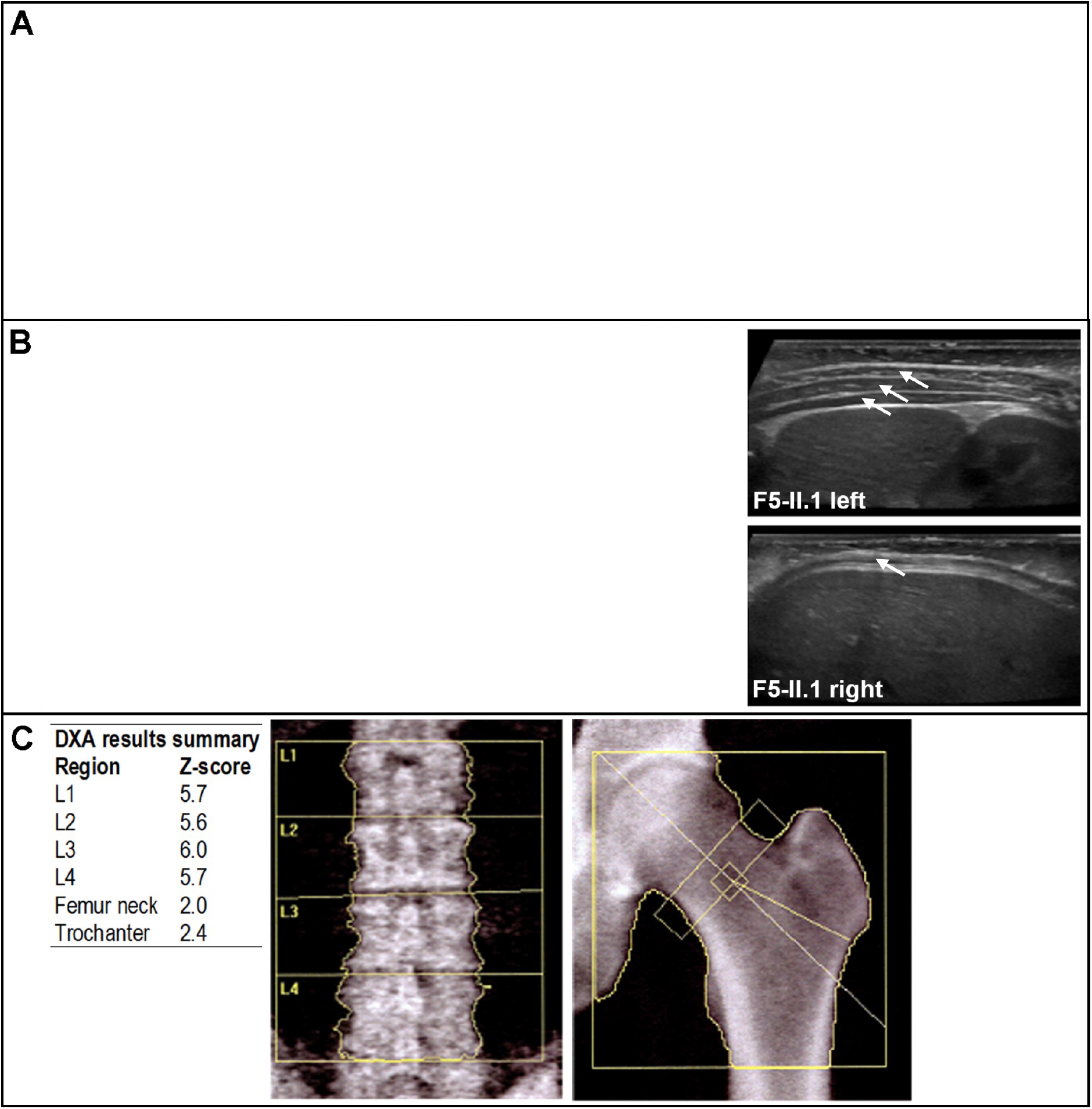
(A) Patient photographs have been removed from preprint version of the manuscript. Please contact the corresponding author for additional information. (B) Ultrasound images demonstrating congenital hypoplasia of the abdominal wall musculature on the right side in the affected male from family 5 (compare arrows showing three normal muscle layers on left and only one muscle layer on right) (C) Bone densitometry performed for an affected adult male from family 1 (F1-III.2). Dual-energy x-ray absorptiometry (DXA) measured on left hip and lumbar vertebrae, showing increased bone mineral density.

**Family 2** is a kindred with two affected males that both died in the neonatal period with severe malformations that included left sided posterior CDH in one and complete absence of the left hemidiaphragm in the other, plus a large anterior body wall defect in both. The body wall defect consisted of supraumbilical muscle deficiency in both cases, and more extensive clefting of the sternum in individual F2-II.1 (Data not shown). Hypoplasia of the corpus callosum was noted at autopsy in individual II.1 but was absent in II.2. The female carrier (F2-I.2) was born with an umbilical hernia that resolved over time without surgical repair, and then subsequently recurred as an epigastric abdominal wall defect following pregnancy. She did not have the same facial features identified in the adult females in family 1, and her inter-pupillary distance is just above the mean for individuals of European ancestry. Chromosome X-inactivation studies were performed in a peripheral blood sample from the female using HpaII digestion at a polymorphic locus in the AR gene and showed a normal (random) inactivation pattern.

**Family 3** has three affected males, all of whom had left-sided posterolateral diaphragm defects. One affected male (F3-IV.2) underwent additional surgical repair of a ureteral abnormality causing hydronephrosis. Additional anomalies, including body wall defects, or dysmorphic features were not reported in the affected individuals, and neurodevelopmental outcomes for the survivors are not available.

**Family 4** has three affected male individuals. Beyond reported surgical repair for CDH, limited phenotypic information is available for the proband (F4-III.3), or for his affected male relatives (F4-III.1 and F4-II.1), who are both deceased. Neurodevelopmental or cognitive concerns were not reported for the proband. The female obligate carriers and are not known to be affected by diaphragmatic defects or any other structural birth defect.

**Families 5 and 6** each contain one affected male and were identified by screening WGS and WES data from a cohort of 735 patients with CDH and their unaffected parents ^7^. The proband in **Family 5** is a male who presented with right-sided CDH and outpouching of the right side of the abdominal wall, which was evident from birth. Abdominal ultrasound showed congenital absence of the internal oblique and transversus abdominis muscles (Figure 2B). The proband in **Family 6** is a male with a diaphragmatic eventration that required surgical repair, and later re-herniation, and no reported body wall defects. Dysmorphic facial features and significant developmental delays were not noted in either of these probands, and there was also no family history of CDH or other birth defects.

The male proband in **Family 7** (F7-II.2) underwent repair of bilateral ventral diaphragmatic hernias, both covered by a hernia sac. He also had a large skin-covered epigastric abdominal wall defect that was surgically repaired. History was also notable for a membranous ventricular septal defect, atrial septal defect, bilateral hydronephrosis, right undescended testis, right inguinal hernia. Dysmorphisms noted at birth included a prominent forehead, hypertelorism, down-slanting palpebral fissures, a broad flattened nasal bridge, anteverted nares, low set ears, micrognathia, a sacral dimple with a hair tuft, and hypoplastic first toenails bilaterally. There were no neurodevelopmental concerns.

**Family 8** is a family with 3 affected males. The proband (F8-II.5) had a left-sided posterolateral CDH covered by a hernia sac that was surgically repaired, and a neurodevelopmental disorder. Other notable findings included a two-vessel umbilical cord, left-sided grade 3 vesicoureteral reflux, intermittent horizontal nystagmus, corneal pannus, and facial dysmorphisms including hypertelorism, down slanting palpebral fissures, low set right ear, short nose with wide nasal root, widely spaced teeth, and a high arched palate. He had delays in speech development and was later diagnosed with autism spectrum disorder and intellectual disability and complex partial seizures. A brain MRI showed enlarged lateral ventricles. The proband had two male relatives who died in infancy from left-sided CDH (F8-II.2 and F8-II.3), though samples from these individuals were not available for genetic testing. The family history is also notable for other individuals with neurodevelopmental disorders.

### Genetic studies

Linkage analysis was performed on members of family 1. A logarithm of the odds (LOD) score above 3, indicating positive association with diaphragmatic defects, was obtained for a 3 Mb region on chromosome band Xq23 between chrX:113,889,739-116,906,942 (hg19) (Supplemental Materials). The region includes *PLS3*, as well as 8 other protein-coding genes and several non-coding transcripts.

Whole exome sequencing (WES) (families 1-4, families 7-8) or whole genome sequencing (WGS) (families 5-6) identified novel missense sequence variants in *PLS3* that were either maternally-inherited (7 families) or *de novo* (in family 7) (Table 2). The *PLS3* variants segregated with the diaphragmatic phenotype and were notably absent from unaffected male relatives of probands in the familial cases. All variants were validated by Sanger sequencing. Significant chromosome anomalies and copy number variants were excluded in all probands with clinical or research chromosome microarray analysis. Except for family 8, no other rare coding candidate variants segregating with the affected individuals were identified. The proband in family 8 carried two additional coding variants of uncertain significance in addition to the *PLS3* variant: a non-maternally inherited c.G52C, (p.G18R) variant in *SMARCA2*, and a maternally inherited c.G133A, (p.G45S) in *SYP*. A paternal sample was not available for sequencing in this family. *SMARCA2* and *SYP* are both linked with neurodevelopmental phenotypes^29,30^, so it is possible that these variants may be contributing to the neurological features in this individual, which are more severe than in the other patients in this report. However, neither *SMARCA2* nor *SYP* have been linked with diaphragm or body wall defects, and therefore, in combination with the family history consistent with X-linked inheritance, the *PLS3* variant is most likely causative.

**TAble 2:**
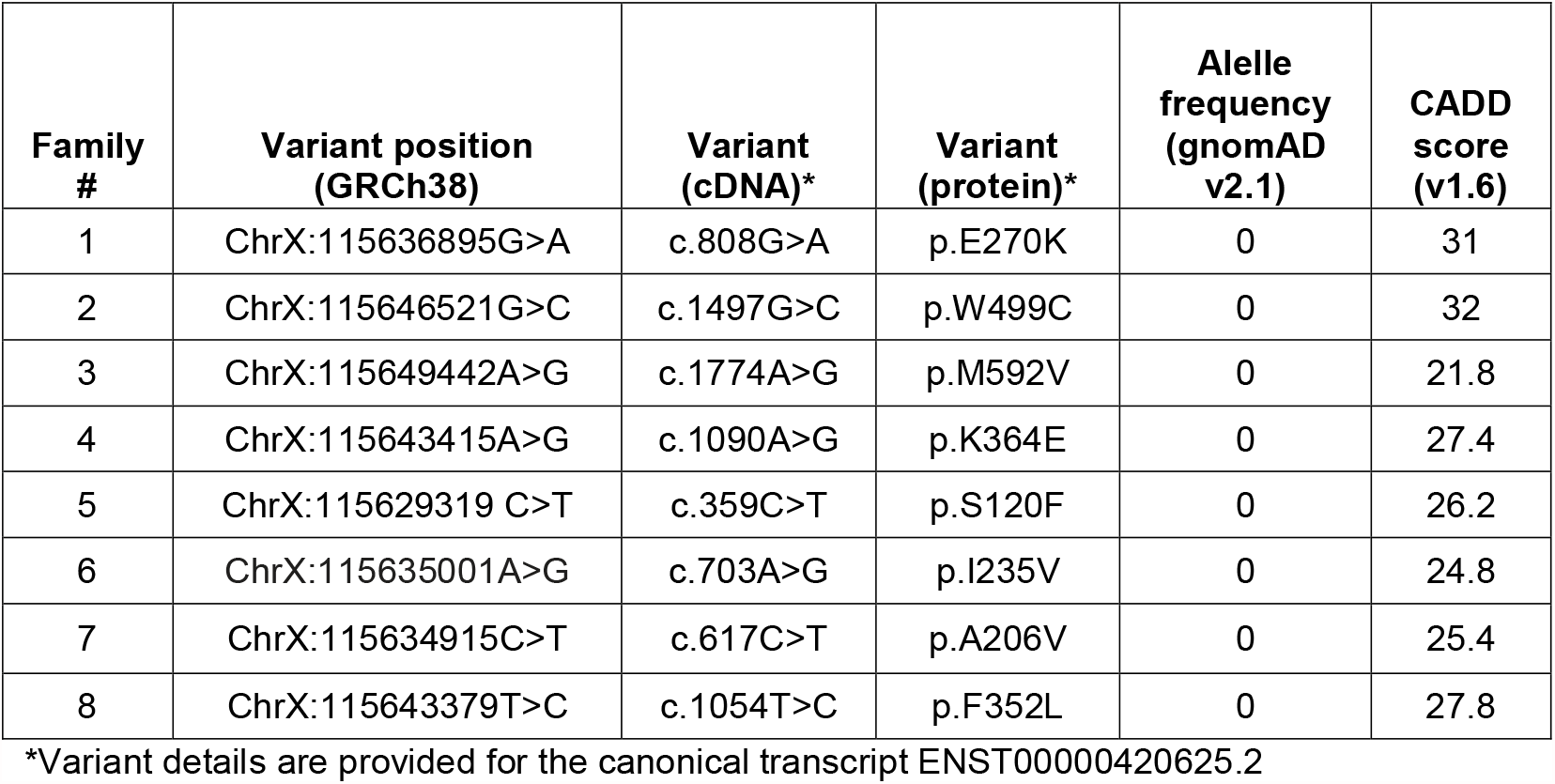
CDH-associated *PLS3* Variant Details.

All eight *PLS3* variants were absent from large population databses (e.g. gnomad^24^), altered conserved residues, and are predicted to be pathogenic by in silico algorithms (e.g. CADD^31^) (Table 2).

### Protein modeling

We used structural modeling of 3D-interactions of actin-bundling proteins to address the potential effects of the identified missense variants (Figure 3). PLS3 is characterized by an N-terminal headpiece and two C-terminal actin-binding domains (ABD1 and ABD2), formed by tandem pairs of calponin-homology (CH1-CH4) subdomains ^16^. All eight variants identified in families with CDH are in ABD1 or ABD2 and none of them are predicted to cause a conformational change in the protein (Fig 3B). The p.Glu270 and p.Trp499 residues affected in families 1 and 2, respectively, are contained in the core residues involved in actin binding (ABS, acting binding sites) ^32,33^. Although the p.Glu270Lys variant (family 1) is not predicted to induce conformational changes, it does disrupt the bond between Gly270 and Lys236 (Fig 3B, magenta), which has been shown to be necessary for binding with F-actin^25^. The p.Trp499Cys change (family 2) in the ABD2 domain is predicted to affect the hydrophobic core of the actin-binding site (Fig 3B, yellow). The p.Met592 residue (family 3) is also part of the ABD2 domain (Fig 3B, orange); however, it does not map to any of the characterized actin-binding sites and therefore the consequences of the missense p.Met592Val cannot be inferred by structural modeling alone. The p.Lys364Glu variant (family 4) is predicted to cause loss of the hydrogen bond between Lys364 and the Asp125 residue located in the actin-binding site of ABD1 (Fig 3B, green). The p.Ser120Phe (family 5), p. Iso235Val (family 6), p.Ala206Val (family 7), and p.Phe352Leu (family 8) variants are all located at the actin binding interface of ABD1(Fig 3B) and, given their locations, may alter actin binding.

**FIGURE 3.**
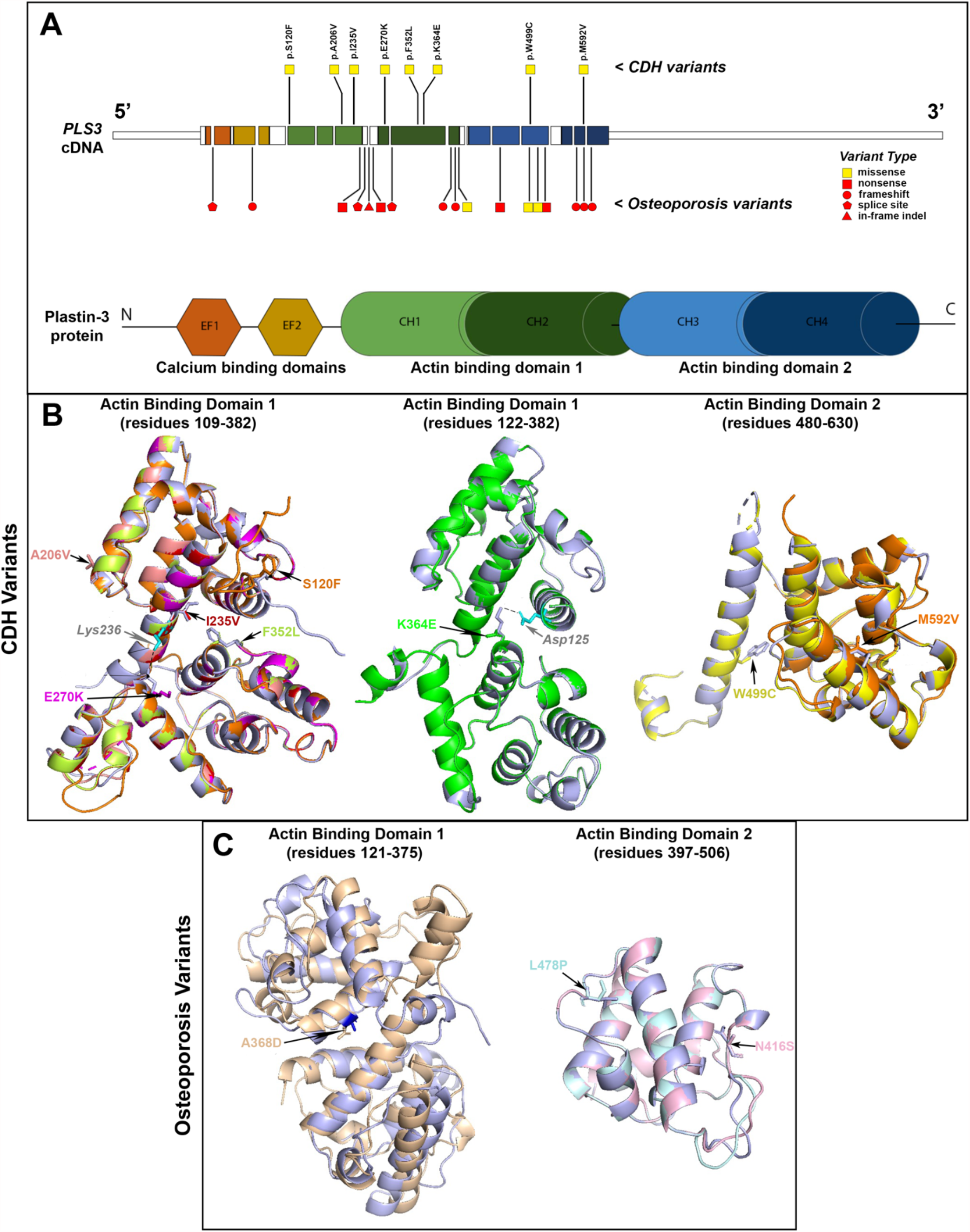
(A) Locations and types of PLS3 variants in patients with CDH and X-linked osteoporosis. The cDNA structure for the canonical PLS3 transcript ENST00000420625.2 is shown in the top panel and the exons are color-coded depicting the corresponding domains in the Plastin-3 protein, shown in the bottom panel. The CDH variants are depicted above the cDNA and the osteoporosis variants are depicted below the cDNA. Yellow squares indicate missense variants, and red shapes indicate loss-of-function variants (red square: nonsense, red circle: frameshift, red pentagon: essential splice site, red triangle: in-frame indel). The Plastin-3 protein image shows the following domain types: EF, EF-hand domain; CH, Calponin-homology domain. (B) Predicted protein structures for the actin binding domains of CDH-associated variants. In these images, the predicted structures of each of the eight variant Plastin-3 proteins (multiple colors) are overlaid on the structure of the native (wildtype) Plastin-3 protein (lavender). The amino acid alterations are written in the same color as the corresponding protein structure. The overlaid structures demonstrate that none of the CDH variants are predicted to cause a major alteration in protein structure. The dotted lines indicate salt bridges between Lys236 and Glu270 (left panel) and Asp125 and Lys364 (middle panel) of the wildtype protein. (C) Predicted protein structures of the actin binding domains of osteoporosis-associated missense variants. The predicted structures of three Plastin-3 variants (peach, pale blue, pale pink) are overlaid on the structure of the native protein (lavender) showing that the osteoporosis variants are predicted to cause a major change in protein structure.

Structural modeling was also performed on the three missense variants previously linked with osteoporosis^34–36^. The p.Ala368Leu alters the conformation of the ABD1 with a high deviation value (3,325Å) (Fig 3C, peach). The p.Leu478Pro variant is predicted to be responsible for major alteration of ABD2 structure since the proline substitution precludes hydrogen bonding with the adjacent amino acid (Fig 3C, pale blue). Finally, the Asn446Ser substitution, also in ABD2, shows the same topology and the same conformation (Fig 3C, pale pink). However, conformation changes can be obscured by the fact that the predicted model is not built with the native structure but with a homologous model having a sequence identity superior to 30%. These findings support a model in which osteoporosis-associated missense variants may be responsible for loss-of-function by major alterations in protein structure, while CDH-associated missense variants do not alter protein conformation but are instead responsible for specific alteration of actin-binding.

### PLS3 overexpression studies

To study the functional effect of a subset of CDH-associated *PLS3* variants on the actin cytoskeleton, we performed overexpression experiments of wild type and variant PLS3 in RFL-6 cells, which are derived from rat embryonic lung mesenchyme and express very low levels of endogenous *Pls3* (data not shown). N-terminal HA-tagged human wild type and mutant *PLS3* constructs were electroporated into RFL-6 cells. Immunofluorescent staining of the HA-tagged PLS3 proteins revealed that all variants are translated and co-localize with phalloidin-stained F-actin in the cellular subcortical region, lamellipodia, and filopodia (Figure 4A). Cells overexpressing all CDH-associated HA-PLS3 variants displayed grossly abnormal morphology compared to cells expressing wild type HA-PLS3, with increased microspikes in the lamellipodia and occasional abnormally long filopodia, as well as frequent abnormal intracellular aggregates of actin (Figure 4A). Cells overexpressing N-terminal HA-tagged Glu270Lys-*PLS3* and Trp499Cys-*PLS3* also showed a quantifiable decrease in the formation of actin stress fibers; furthermore, this effect was not seen by overexpressing the osteoporosis-associated missense variant Leu478Pro-*PLS3*^34^ (Figure 4B-C). An effect on cell migration was hypothesized but could not be quantified easily using this cell model. These experiments demonstrate that CDH-associated variants in the actin binding domains of PLS3 perturb the actin cytoskeleton, and that these effects differ from those caused by an osteoporosis-associated missense variant.

**FIGURE 4.**
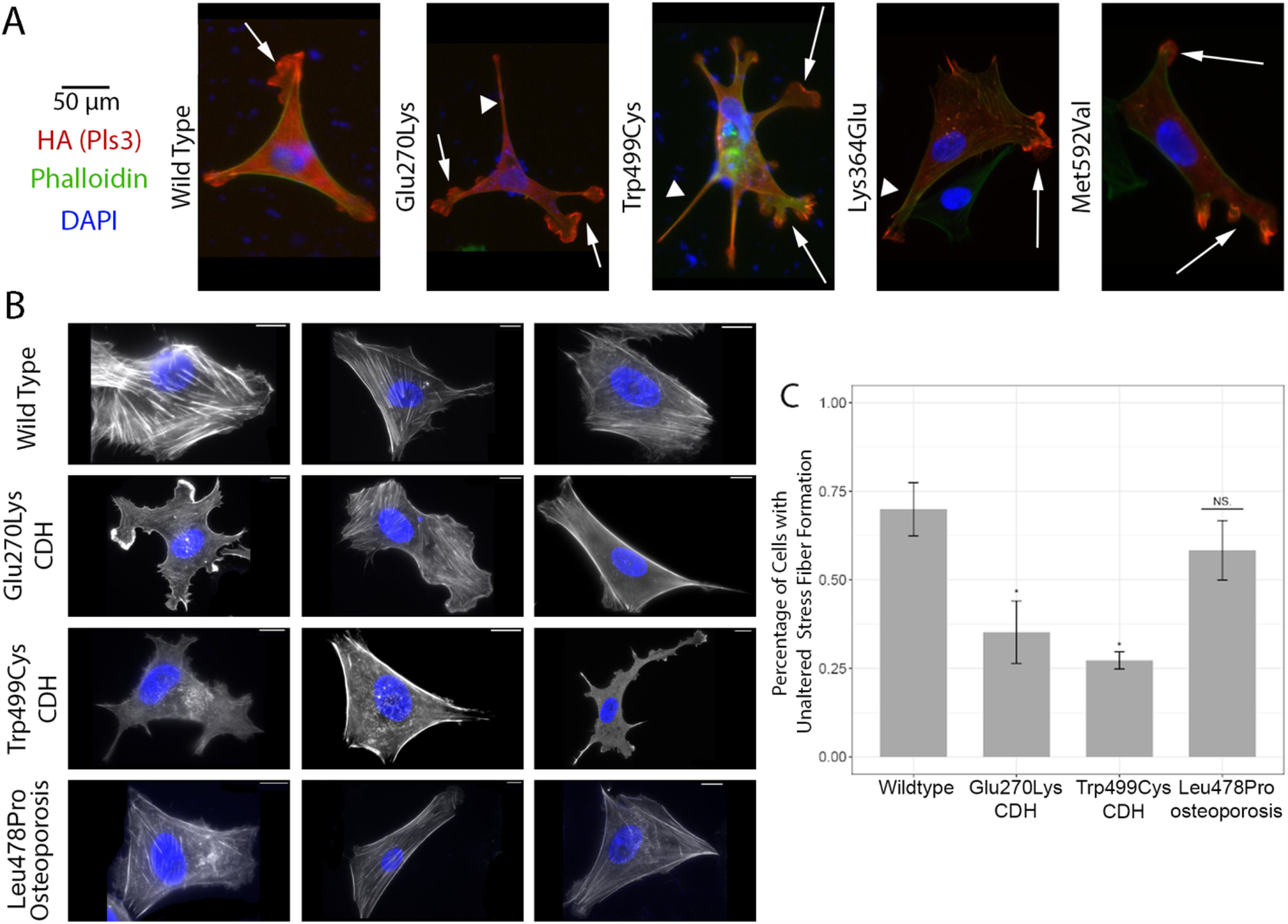
(A) Overexpression of HA-tagged wild-type and variant PLS3 in RFL-6 cells. Green: fluorescein-conjugated phalloidin, staining for filamentous actin. Red: anti-HA antibody, staining for exogenous HA-PLS3 protein. Left panel: wild-type PLS3 is distributed homogeneously in RFL-6 embryonic lung fibroblasts (red); actin fibers are normally aligned in the cytoplasm (green). Right panels: mutant PLS3 accumulates on the cell edges where it is strongly associated with F-actin; actin fibers are abnormally positioned, resulting in distorted cell morphology. Additionally, mutant PLS3 is highly expressed in abnormal and increased number of lamellipodia (arrows) and filopodia (arrowheads). (B) Phalloidin staining (white) to visualize actin stress fibers in RFL-6 cells overexpressing wildtype PLS3, two CDH-associated Pls3 variants (Glu270Lys and Trp499Cys) and one osteoporosis-associated PLS3 missense variant (Leu478Pro). (C) The percentage of cells in the experiment depicted in (B) showing normal actin stress fiber formation was quantified in a blinded analysis as described in the methods. ^*^ p-value <0.001.

### Mouse model of the PLS3 W499C variant

We investigated the expression pattern of *Pls3* mRNA in mouse embryos during critical times of diaphragm, lung, and body wall development. PLS3 has been described as being expressed almost ubiquitously in adult non-hematopoietic cells with replicative potential^37^. However, in situ hybridizations showed a complex staining pattern in mouse embryos, demonstrating that this gene is regulated in a tissue-specific manner during embryonic development (Fig 5A). At E12.5 and E13.5, *Pls3* is expressed in the diaphragm tissue, between liver and heart, as well as the lung (Figure 5B,C). Strong staining was observed in the lung mesenchyme at E13.5 (Fig 5D). PLS3 protein expression was also present in the mesenchyme and alveolar septae of human neonatal lung specimens (Supplemental Materials). Finally, we detected strong expression in lateral paired and symmetric structures representing the growing edges of the body wall in mouse (Fig 5B), pertinent to the body wall defects seen in some patients.

**FIGURE 5.**
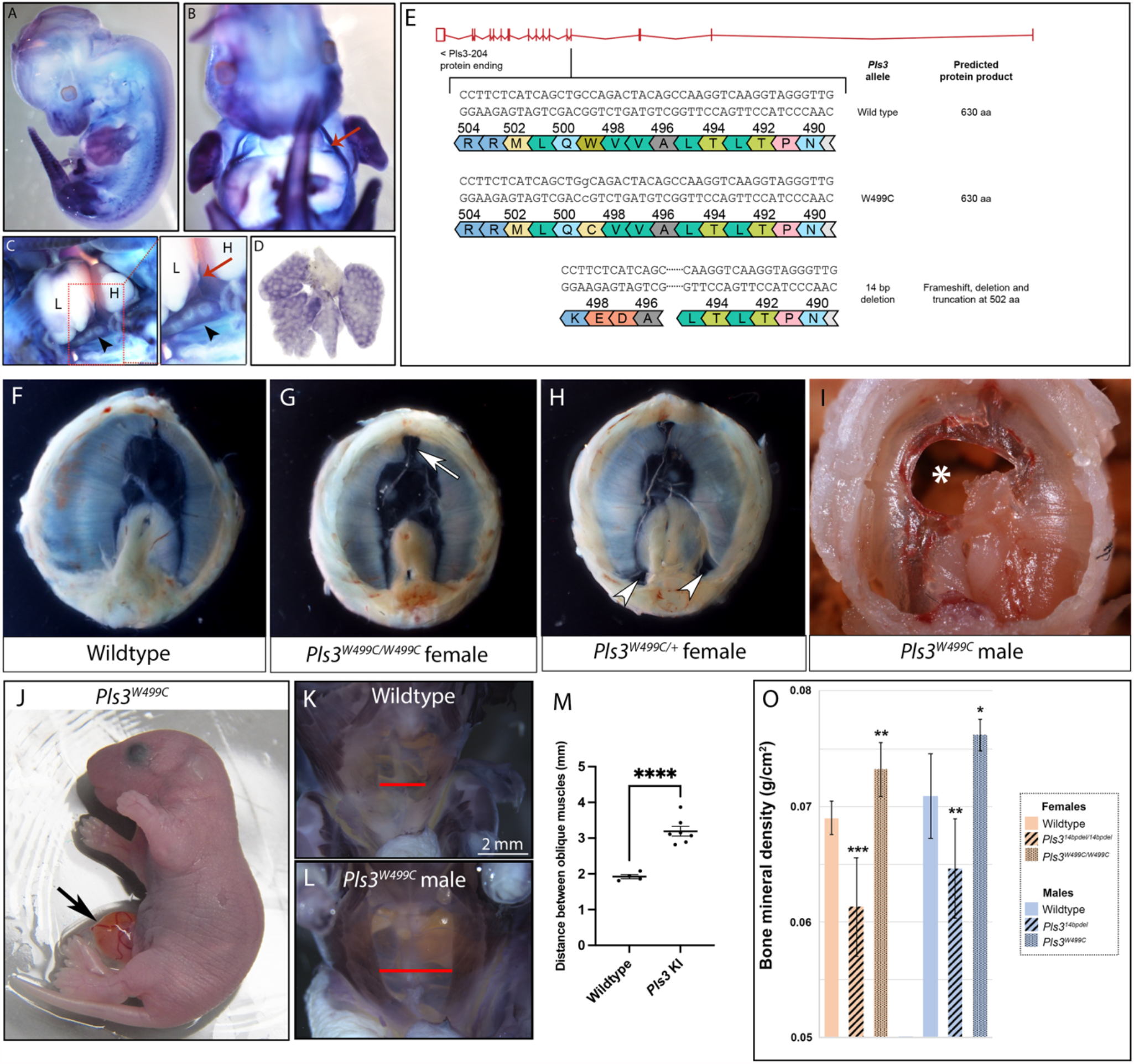
(A-D) Whole-mount *in situ* hybridization for Pls3 in mouse embryos at E12.5 (A-C) and E13.5(D). (A) Pls3 mRNA is observed in face, limbs, tail and axial skeleton in a E12.5. (B) The ventral view shows Pls3 in symmetrical structures lateral to the midline (arrow) with a distribution compatible with edges of the anterior body wall. (C) Dissection of the lateral body wall shows that Pls3 mRNA is expressed in the developing lung (arrowhead) but not in the heart (H) or in the liver (L). The developing diaphragm, nested between the heart and the liver, shows Pls3 expression (red arrow in insert). (D) Microdissection of the lung shows Pls3 expression in the lung is specific to the mesenchyme at E13.5. (E-F). (E) Targeting strategy for Pls3W499C knockin and Pls314bpdel mice. The knock-in results in a single amino acid substitution at position 499. The 14bp deletion results in a deletion and frameshift, with predicted truncation of the protein at 502aa. (F-I) Images of the diaphragms dissected from E18.5 or P0 wildtype and Pls3W499C mutant mice. Abnormalities seen in the mutants include muscular thinning anteriorly under the sternum (arrow in G), muscular thinning posterolaterally (arrows in H) and complete holes in the diaphragm (^*^ in I). (J) Omphalocele (arrow) in a Pls3W499C mutant at P0. (K-L) Whole mount myosin staining to visualize the body wall musculature in wildtype and Pls3W499C mutants at P0. Red bars show the distance between the external oblique muscles. (M) Quantification of the distance between the external oblique muscles showing a statistically significant increase in the Pls3W499C knock-in compared to controls. ^****^ represents P<0.0001. Error bars on graph represent the standard error of the mean, centered on the mean of each genotype. (O) Bone densitometry studies of 3 month-old Pls314bpdel and Pls3W499C mice compared with age- and sex-matched controls. Bone mineral density values are shown for wildtype females (n=8), Pls314bpdel homozygous females (n=7), Pls3W499C homozygous females (n=6), wildtype males (n=8), Pls314bpdel hemizygous males (n=8), and Pls3W499C hemizygous males (n=3). P-values compared with sex-matched controls: ^*^ <0.05, ^**^ <0.002, ^***^ <0.0005. Error bars indicate one standard deviation of the mean.

To model the effect of one human CDH-associated variant *in vivo*, we generated a knock-in mouse model, *Pls3*^*W499C*^, carrying the W499C variant observed in Family 2. Another line, *Pls3*^*14bpdel*^, which is predicted to cause a frameshift, was also generated from the same germline targeting experiment (Fig 5E). The *Pls3*^*14bpdel*^ mice were therefore used as a loss-of-function comparison with the *Pls3*^*W499C*^ mice.

*Pls3*^*W499C*^ mice were found at expected genotype ratios at late gestation, but there was a significant reduction in the number of hemizygous males and homozygous females that survived to weaning, and most affected pups died within the first 2 days after birth. In contrast, the *Pls3*^*14bpdel*^ mice were viable and fertile, consistent with another published *Pls3* knock-out model^38^. The *Pls3*^*W499C*^ mutants displayed several phenotypes at late gestation and neonatal time points, including diaphragm abnormalities and anterior body wall defects, both with variable penetrance. The diaphragm defects included thinning of the muscular diaphragm, particularly at the posterolateral edge and the anterior region below the sternum.

Less common complete holes in the diaphragm were also observed (Fig 5G-I, Table 3). The abdominal wall defects included midline protrusion of the intestine into a sac at the site of the umbilicus, similar to omphalocele, in a subset of mutant animals (Fig 5J). All the mutants showed thinning of the abdominal wall muscles with widening of the space between the external oblique muscles (Fig 5K-M). Neither diaphragm nor abdominal wall defects were observed in the *Pls3*^*14bpdel*^ mice (Table 3, data not shown). We also examined the lungs from *Pls3*^*W499C*^ *mice*, as primary defects of the lung mesenchyme are believed to be linked to CDH-associated pulmonary hypoplasia^39,40^, but we did not observe and major defects in lung structure or cellular markers (supplemental data).

**TAble 3:** Summary of diaphragm phenotypes in Pls3 mouse models.

| Genotype | Sex | n | Percent abnormal | Type of Diaphragm Defect |  |  |
| --- | --- | --- | --- | --- | --- | --- |
|  |  |  |  | Anterior muscular thinning | Posterolateral muscular thinning | Muscular and membranous defect |
| Wildtype | M | 5 | 0% | - | - | - |
| *Pls3*<sup>W499C/+</sup> | F | 6 | 50% | 3/6 | - | - |
| *Pls3*<sup>W499C/W499C</sup> | F | 16 | 87.5% | 14/16 | 6/16 | 1/16 |
| *Pls3*<sup>W499C</sup> | M | 6 | 83.3% | 5/6 | 3/6 | 1/6 |
| *Pls3*<sup>14bpdel/14bpdel</sup> | F | 4 | 0% | - | - | - |
| *Pls3*<sup>14bpdel</sup> | M | 3 | 0% | - | - | - |

To determine the effect of the W499C variant on bone density, we performed DEXA scans on surviving homozygous female and hemizygous male knock-in mice at 3 months of age (Fig 5N). Compared with age- and sex-matched wildtype control mice, the *Pls3*^*W499C*^ knock-in mice showed increased bone mineral density. In contrast, the loss-of-function *Pls3*^*14bpdel*^ mice showed decreased bone mineral density compared to controls (Fig 3N).

## DISCUSSION

The human and mouse data in this study show that missense variants in the actin binding domains of *PLS3* are associated with a congenital disorder characterized by diaphragm defects and variable anterior body wall defects, for which we propose the name “*PLS3*-associated congenital anomaly syndrome”. Affected individuals in three families also showed dysmorphic facial features, with hypertelorism being a common finding. It is therefore possible that this syndrome contains a distinctive facial gestalt, though this will require additional investigation as detailed dysmorphology examinations were not available for the other patients. Neurodevelopmental features were notably absent from most of the surviving individuals, except for two adult males in family 1 and the surviving proband in family 8. In these cases, additional environmental or genetic factors contributing to the neurodevelopmental phenotype could not be ruled out.

The co-occurrence of diaphragm and ventral body wall defects is reminiscent of a pattern of malformation affecting the embryonic midline that has been recognized since the 1950’s, described by others as Pentalogy of Cantrell^41^ and thoracoabdominal syndrome (TAS) ^42,43^. We identified some commonalities and differences between *PLS3*-associated congenital anomaly syndrome and these malformation syndromes. Several minor features of TAS, including cardiac defects, cystic hygroma, and hydronephrosis, were each seen in a minority of the individuals we describe with *PLS3* variants. The diaphragm defects in the patients with *PLS3* variants are predominantly posterolateral, which differ from the ventral diaphragm defects described in most cases of Pentalogy of Cantrell and TAS. While Pentalogy of Cantrell is typically sporadic, TAS has been reported in families consistent with X-linked inheritance^42,43^. Linkage studies in one family mapped a candidate locus to a different region of the X chromosome (Xq27) that does not contain the *PLS3* gene^44^. It is therefore likely that variants in several different genes can result in overlapping patterns of malformations.

The Plastin family of actin bundling proteins have been shown to play multiple diverse cellular roles, including regulation of cell shape, motility, adhesion, endocytosis, vesicle trafficking, and organization of specialized cellular structures such as microvilli. The tissue-specific developmental expression pattern of Pls3 is consistent with a role for this gene in the congenital anomalies observed in the patients in this study. Development of the diaphragm requires complex cellular processes including migration and differentiation of the pleuroperitoneal folds, which form the diaphragm’s connective tissue and central tendon, as well as skeletal muscle precursors derived from the somites^45,46^. Similarly, cell migration processes are likely critical for development of the abdominal wall^47^. Plastin 3 has been hypothesized to be particularly important for promoting the migration of cells across gaps in the extracellular matrix by stabilizing cellular membrane protrusions^48^. Further research into the effects of actin binding domain variants on cellular processes during development should shed additional insight into the mechanism of the congenital anomalies seen in these patients.

Loss-of-function variants in *PLS3* have been described in multiple families to cause X-linked osteoporosis and osteoporotic fractures (MIM# 300910) (Supplemental Materials) ^13^. Loss-of-function of *Pls3* in mouse also recapitulates the human osteoporosis phenotype^38^. Nearly all osteoporosis-related human variants in *PLS3* result in protein truncation or abnormal splicing, and the few missense and in-frame insertion variants in this disorder render the protein either hyporesponsive or hyperresponsive to calcium or completely disrupt actin binding ability *in vitro*^49^. Therefore, loss-of-function or abnormal calcium responsiveness of PLS3 are the major mechanism in osteoporosis, and the results of our *in silico* modeling for three missense osteoporosis variants are consistent with this. In contrast, our data support a model that CDH-associated *PLS3* variants perturb actin binding and actin cytoskeletal dynamics without causing a major change in protein structure. We hypothesize that these specific cellular functions of PLS3 underly the anomalies seen in the patients.

The observation of increased bone mineral density in the *Pls3*^*W499C*^ knock-in mouse model and a patient from family 1 suggests that these variants may have a gain-of-function effect, at least in bone. A gain-of-function model is also supported by data on the homologous yeast protein, fimbrin, encoded by the *SAC6* gene. The yeast sac6 suppressor mutation Trp514Cys (homologous to the p.Trp499Cys variant identified in family 2), results in a stronger interaction with actin compared with wild-type^50^. These yeast data also support the pathogenicity of the human p.Trp499Cys variant, specifically. Overexpression of wildtype Pls3 in mice results in increased bone mineral density but did not result in major congenital anomalies^38^, and approximately 5% of the human population also over-expresses PLS3 without any apparent clinical consequences^13^. Therefore, abnormal plastin-actin interaction, rather than abnormal stoichiometry, is the most likely explanation for the diaphragm and body wall defects. Additional detailed biochemical experiments will be helpful to quantify the differing effects of these classes of human variants on Pls3 function in actin-bundling and calcium responsiveness.

Together, these results show that CDH-associated *PLS3* variants cause a distinct syndrome from the loss-of-function variants identified in patients with osteoporosis, and support the hypothesis that these missense variants may be gain-of-function.

## Data Availability

WES data from families 2-4 has been deposited into the NIH National Center for Biotechnology Information (NCBI) Database of Genome and Phenotypes (dbGAP). WES data from families 7 and 8 will be deposited into NHGRI Genomic Data Science Analysis, Visualization, and Informatics Lab-space (AnVIL). WGS data for families 5 and 6 will be deposited into the data repository of the NICHD Gabriella Miller Kids First (GMKF) Program.

## Acknowledgements

The study was funded by grants from the National Institute of Child Health and Human Development (NICHD/NIH, http://www.nichd.nih.gov) 2P01HD068250 (P.K. Donahoe), R01 HD057036 (W. Chung), and RO1 HD098458 (D. Scott), the National Human Genome Research Institute (NHGRI) and the National Heart Lung and Blood Institute (NHBLI) to the Baylor-Hopkins Center for Mendelian Genomics (BHCMG, UM1 HG006542, J.R. Lupski). Sequencing services were partially provided through the RS&G Service by the Northwest Genomics Center at the University of Washington, Department of Genome Sciences, under U.S. Federal Government contract number HHSN268201100037C from the National Heart, Lung, and Blood Institute (M. Longoni, PI), and from the Gabriella Miller Kids First Pediatric Research Program X01 HL132366, X01 HL136998, and X01 HL140543 (W. Chung, PI).

We are grateful to Eric Liao MD, PhD for important experiments in the zebrafish model and discussion that helped understanding of the different effects of loss-of-function variants in Pls3. Christine Wooley (The Jackson Laboratory Center for Biometric Analysis) performed the DEXA assay on the laboratory mice.

